# Assessing the validity of dual-minima algorithm for heel-strike and toe-off prediction for the amputee population

**DOI:** 10.1101/2021.08.27.21262720

**Authors:** Zohaib Aftab

## Abstract

Assessment of gait deficits relies on accurate gait segmentation based on the key gait events of heel strike (HS) and toe-off (TO). Kinematics-based estimation of gait events has shown promise in this regard especially using the leg velocity signal and gyroscopic sensors. However, its validation for the amputee population is not established in the literature. The goal of this study is to assess the accuracy of lower-leg angular velocity signal in determining the TO and HS instants for the amputee population. An open data set containing marker data of 10 subjects with unilateral transfemoral amputation during treadmill walking was used. A rule-based dual-minima algorithm was developed to detect the landmarks in the shank velocity signal indicating TO and HS events. The predictions were compared against the force platform data for 2595 walking cycles from 239 walking trials. Results showed considerable accuracy for the HS with a median error of -1ms. The TO prediction error was larger with the median ranging from 35-84ms. The algorithm consistently predicted the TO earlier than the actual event. Significant differences were found between the prediction accuracy for the sound and prosthetic legs. The prediction accuracy was also affected by the subjects’ mobility level (K-level) but was largely unaffected by gait speed. In conclusion, the leg velocity profile during walking can predict the heel-strike and toe-off events for the transfemoral amputee population with varying degrees of accuracy depending upon the leg side and amputee’s functional ability level.

## 1 Introduction

Gait analysis is a valuable tool in assessing various pathologies. Deviations from a healthy gait template often indicate underlying health conditions. For people with amputations, an accurate gait assessment leads to the development of more efficient prosthetic devices and better evaluation of clinical outcomes. That is why, more and more clinics are using technology-enabled gait analysis solutions as opposed to traditional visual gait analysis.

A pre-requisite to effective gait assessment is the estimation of key gait events of heel strike (HS) and toe-off (TO), which represent the moment the foot is placed and removed from the ground respectively [1]. Traditionally, these are estimated using the force or pressure data from specialized platforms in research laboratories. However, due to high cost and space constraints, this method is not used widely in clinics. Instead, kinematics-based solutions are becoming popular due to the possibility of body-worn inertial sensors. These methods rely on leg or foot velocity/acceleration data and rule-based algorithms to estimate gait events. Many authors have validated this approach for TO and HS detection for healthy subjects [2, 3, 4, 5, 6, 7, 8, 9, 10, 11] as well as for subjects with walking disorders [12, 13, 14, 15, 16]. However, its validity for the amputee population is not well established in the literature.

The kinematic methods require an algorithm to identify observable features in the velocity/acceleration data of body segments. Several rule-based algorithms have also been developed for this purpose. A popular choice is to use the shank angular velocity for TO and HS estimation corresponding to the minima in the sagittal-plane angular velocity signal [17]. Many researchers have exploited this signal over the years for diverse subject populations and reported a reasonable degree of accuracy [18, 19, 7, 3, 20, 14, 21, 22, 2].

However, no study has focused on amputee subjects except [20] which included data from a single subject. Since lower-body amputations lead to gait deviations and compensatory movements, there is a need to evaluate this paradigm with a large data set of persons with amputations. Hence, the goal of this study is to compare the accuracy of gait event prediction using the shank angular velocity against force platform data. A secondary objective is to observe if this accuracy is affected by parameters such as leg side (sound or prosthetic), subjects’ walking ability, and walking speed. A published data set of amputee subjects by Hood et. al. [23] is employed for this purpose containing optoelectronic and force plate data for transfemoral amputees.

## 2 Materials and Methods

A data set consisting of marker and force plate data of 18 individuals with unilateral transfemoral amputation was recently published [23]. It is the most comprehensive gait data set available for prosthesis users which provides force platform data for all steps taken during a trial. Moreover, the walking speed was controlled accurately on a treadmill as opposed to subjective instructions to walk ‘slow’ or ‘fast’.

Subjects were divided into two groups based on their comfortable walking speed with the speed of 0.8m/s acting as a threshold. On the Medicare functional classification level (MFCL), subjects were either categorized as limited community ambulators (K-level 2) or full community ambulators (K-level 3) [24]. Each subject walked at five different speeds with K-level 2 subjects at [0.4, 0.5, 0.6, 0.7, 0.8m/s], and K-level 3 subjects at [0.6, 0.8, 1.0, 1.2, 1.4 m/s]. Complete details on the protocol and data acquisition are available in [23].

The original study contained an equal number of subjects in both groups. However, for this study, subjects using the handrails during walking were excluded to avoid the effect of secondary support on the gait pattern. This resulted in ten subjects for further analysis (including three K-level 2 and seven K-level 3 subjects). These subjects are listed in Table 1.

**Table 1:**
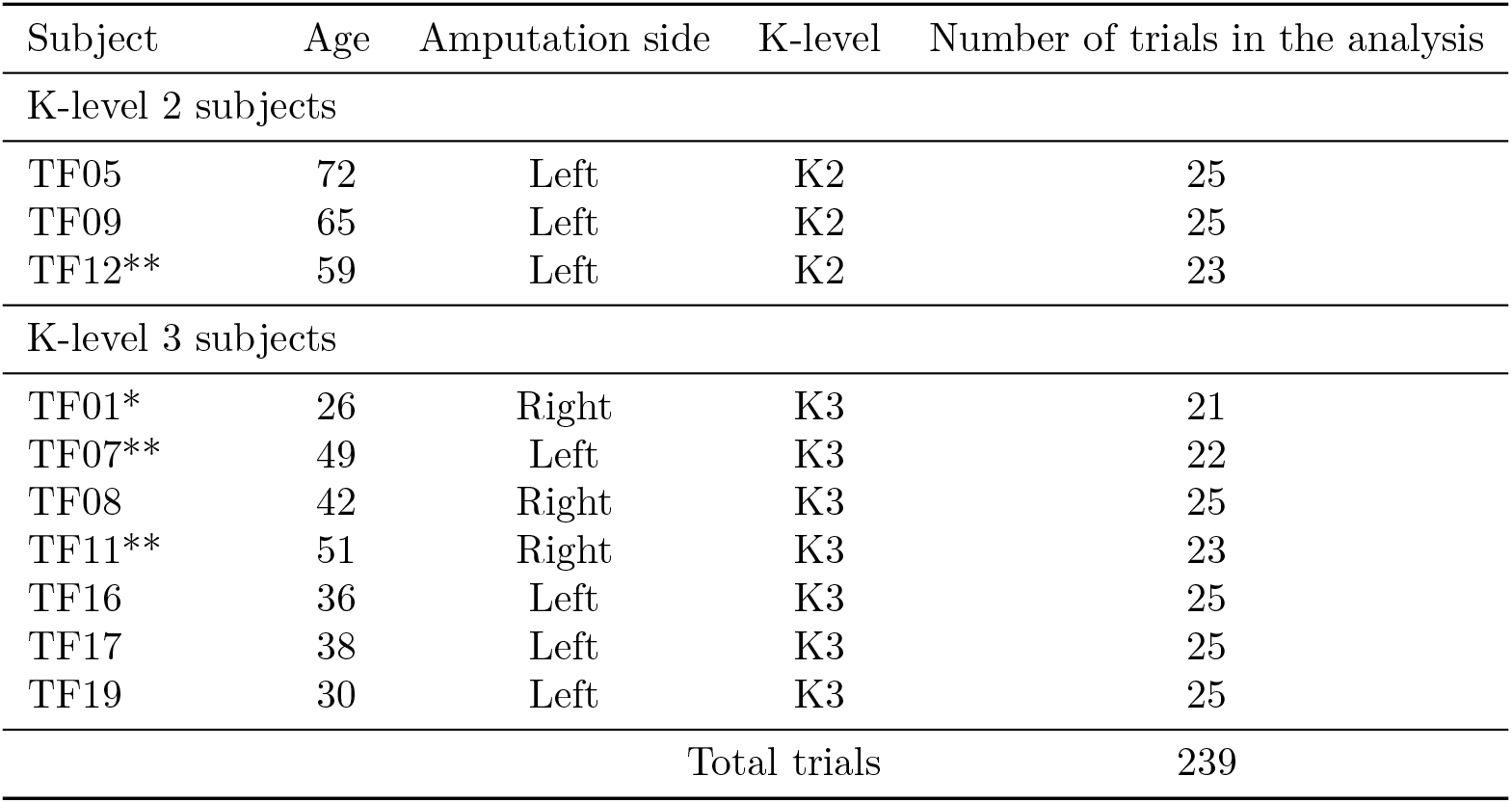
List of subjects for whom the walking data is used in this study. Complete details on amputation can be found in [23]

| Subject | Age | Amputation side | K-level | Number of trials in the analysis |
| --- | --- | --- | --- | --- |
| K-level 2 subjects |  |  |  |  |
| TF05 | 72 | Left | K2 | 25 |
| TF09 | 65 | Left | K2 | 25 |
| TF12** | 59 | Left | K2 | 23 |
| K-level 3 subjects |  |  |  |  |
| TF01* | 26 | Right | K3 | 21 |
| TF07** | 49 | Left | K3 | 22 |
| TF08 | 42 | Right | K3 | 25 |
| TF11** | 51 | Right | K3 | 23 |
| TF16 | 36 | Left | K3 | 25 |
| TF17 | 38 | Left | K3 | 25 |
| TF19 | 30 | Left | K3 | 25 |
| Total trials |  |  |  | 239 |

The data set reported four to five walking trials per speed resulting in a total of 246 walking trials for the ten subjects. However, after careful observation of the force platform and marker profiles, some trials were discarded due to either incomplete or erroneous data. This resulted in a total of 239 trials for the final analysis.

### Estimation of leg velocity signal from marker data

The raw data consisted of three-dimensional trajectories of 61 cutaneous reflective markers. The data contained the .c3d files for the marker trajectories which were extracted using an open-source motion analyzer software MOKKA (Motion Kinematic and Kinetic Analyzer [25]). For each trial, the data were exported into a .csv file and read in Matlab for the calculation of lower-leg angular velocities from the coordinates of tibia markers. Two tibia markers on each leg (Figure 1) were used to estimate leg orientation using the method presented in [26]. The method uses two markers in line with the bone axis to calculate the orientation of the segment, which is further differentiated with respect to time to obtain the angular velocity.

**Figure 1:**
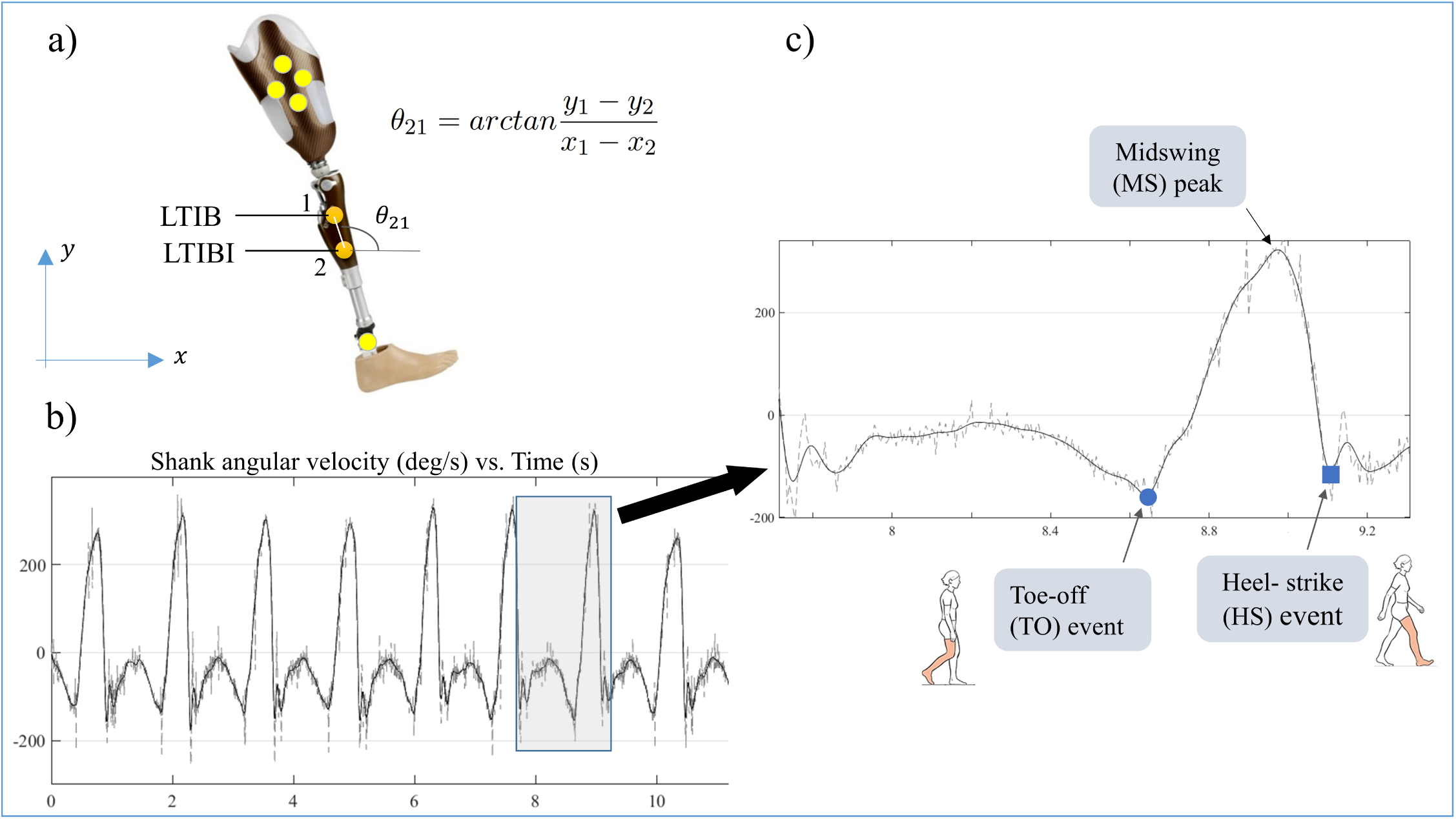
a): Placement of tibia markers for leg orientation and velocity calculation, b) A typical shank velocity signal with raw (grey) and filtered (black) data, c) Enlarged view of one gait cycle from the velocity signal. The algorithm starts with the detection of the largest positive peaks in the signal (marked as MS) which define the intervals for gait events. TO is identified as the last negative peak (or minima) just before the MS (circle) while HS is defined as the negative peak just after the MS (square)

The raw marker data was collected at 200Hz and it is subject to a lot of noise due to soft tissue artifacts. To reduce the noise in the resulting angular velocity signal, a low pass filter was designed and implemented. For this purpose, the frequency spectrum and the Nyquist frequency of the signal for all subjects were analyzed. A cut-off frequency of 4 Hz gave was chosen which resulted in negligible loss of data and time-shift of the signal. An example of the filtering is shown in Figure 1 (right panel) for a trial at 1.2*m.s*^*−*1^.

### Algorithm

The determination of TO and HS events in the velocity signal is based on the dual-minima approach similar to the one presented by [22]. It starts with the detection of all the positive peaks of the signal. These positive peaks are associated with the midswing (MS) (c.f. Fig. 1). Each positive peak is accompanied by two negative peaks (or minima) on either side which indicate the reversal of leg velocity direction. The negative peak (NP) preceding the MS is identified as the toe-off event while the NP after the MS is marked as heel strike. The algorithm is implemented in Matlab.

### Statistical analysis

For each walking cycle, the timings for the TO and HS events obtained by this algorithm are compared against the force platform-based timings provided in the data set. The errors (eTO, eHS) are calculated by taking the difference between the corresponding predicted and actual events.

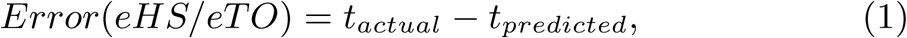

where the actual events refer to the ones marked using the force platform. The error is positive when the predicted event precedes the actual event and vice versa.

Shapiro-Wilk test and visual inspection of histograms indicated that the error distribution was not normal. Hence a 5 number summary statistic (involving the median, lower and upper quartiles, minimum and maximum values) was selected for further descriptive analysis. Non-parametric statistical tests of significance were performed (at p = .05 level) for group differences.

Descriptive statistics of mean error (ME) and mean absolute error (MAE) were also computed to compare the results of this study with the literature which frequently report these averages.

## 3 Results

TO and HS events for a total of 2595 walking cycles from 239 trials were compared. Results are summarized in Figure 2. The median heel strike error (eHS) was -1ms with an interquartile range (IQR) of 31ms. The box extended both above and below the median, indicating the predictions to be early or late than the actual event. On the other hand, the median toe-off error (eTO), as well as the IQR, were larger (43ms and 51ms respectively). The error is positive in all the cases indicating consistent early detection by the algorithm for TO. The mean absolute values shown at the bottom of the plot indicate that the magnitude of eHS was half as compared to eTO indicating a better HS estimation.

**Figure 2:**
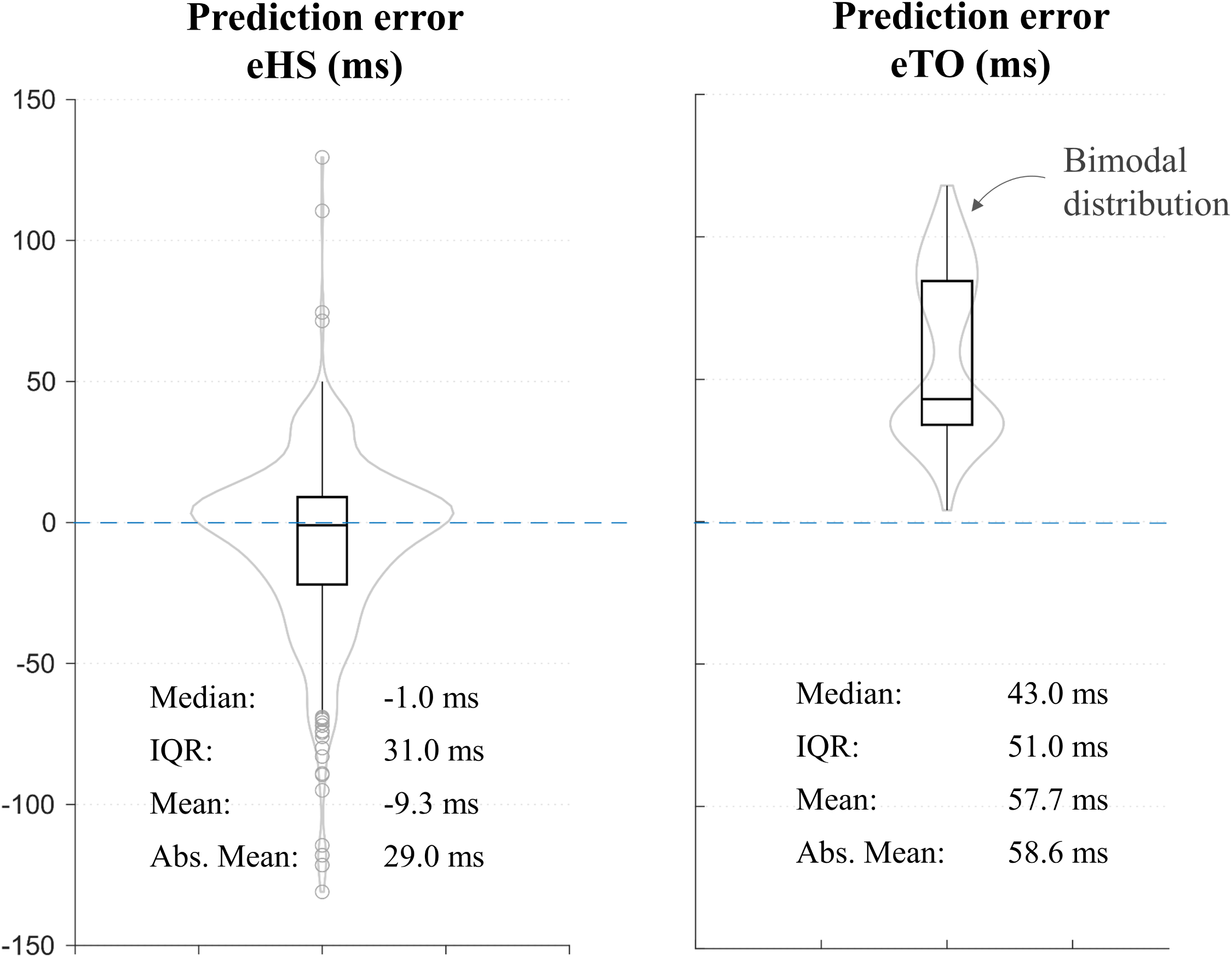
Heel strike (left) and Toe-off (right) error plots for all 2595 walking cycles. Boxplots (black) are superimposed with violin plots (in grey) indicating error distribution. Key statistics are also indicated below each plot. The box indicates the lower and upper quartiles with the central line showing the median. The top and bottom lines of the box represent, respectively, the medians for the upper and lower halves of the data and the whiskers represent the highest and lowest values of the distribution, excluding outliers. Outliers are also presented as circles

The accompanying violin plots (in grey) show the distribution of error values for both events. Interestingly, the distribution is bimodal in nature for the eTO with two local maxima. This indicates a dichotomy of results into two groups.

### Difference by the leg side

To observe the differences in prediction error between legs (sound vs. prosthetic), separate error values are plotted for each leg in Figure 3. A Wilcoxon signed-rank test was also performed to reveal any significant differences.

**Figure 3:**
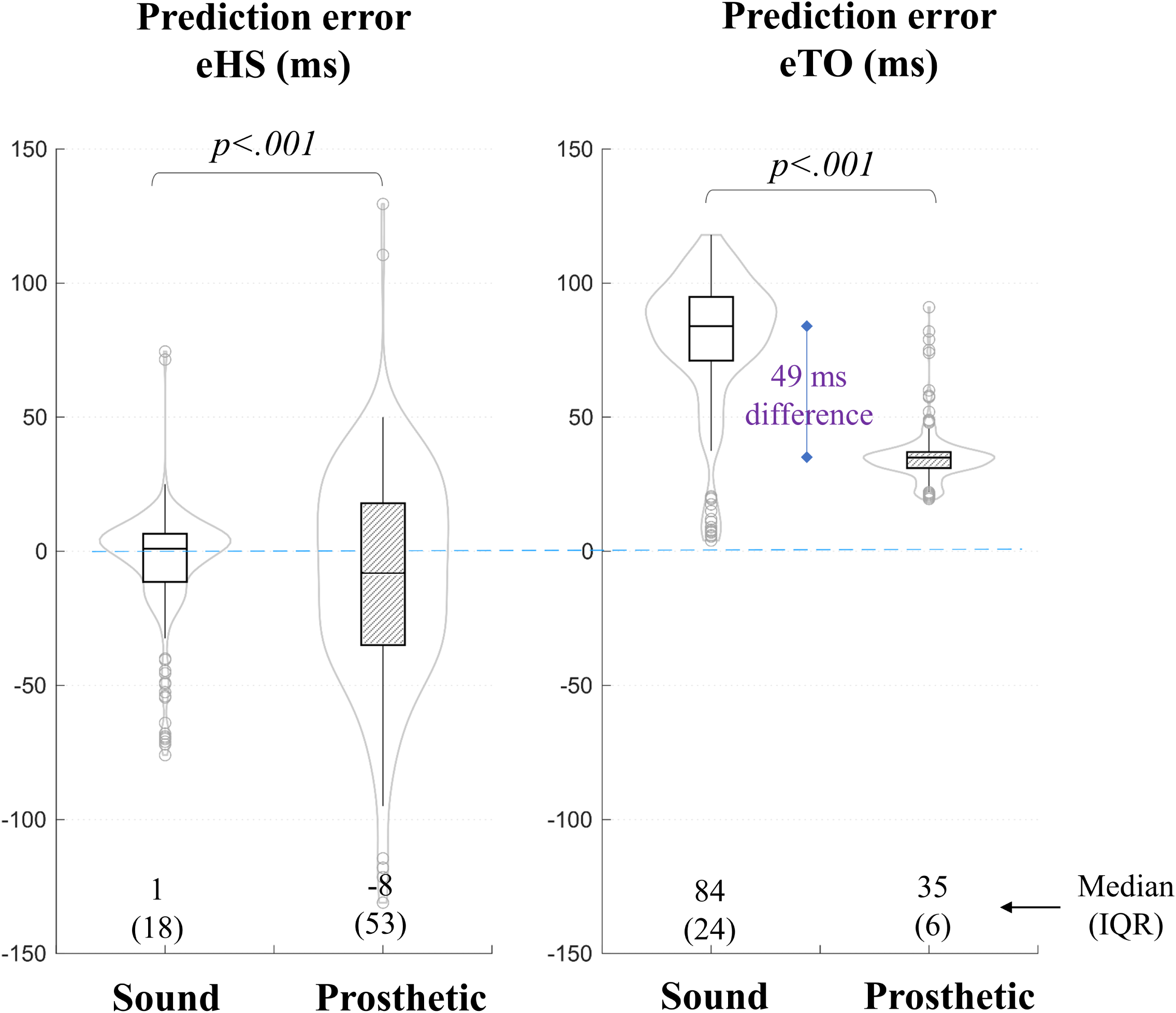
Separate boxplots and error distribution for sound and prosthetic legs. Median and interquartile ranges are mentioned at the bottom in each case. Significant differences were found between errors on the sound and prosthetic sides.

For the heel strike, the median error remained closer for both legs (1ms and -8ms for sound vs. prosthetic side). But the error showed larger dispersion on the prosthetic side with an IQR of 53ms. For the TO event, there was clearly a large difference in the median error value between the two legs. The median error on the sound side was 84ms versus 35ms on the prosthetic side. This explains the bimodal distribution of error observed in Figure 2.

Statistically significant differences were found between the two sides for both TO and HS events (*p <* .*001*) .

### Effect of K-level and walking speed

Finally, in order to observe the effect of subjects’ functional classification level and walking speed on the prediction error, separate boxplots are produced for both subject groups at different walking speeds (Figure 4). Plots for trials from K2 subjects are shown on the left while trials from K3 subjects are on the right. Walking speed is varied on the x-axis.

**Figure 4:**
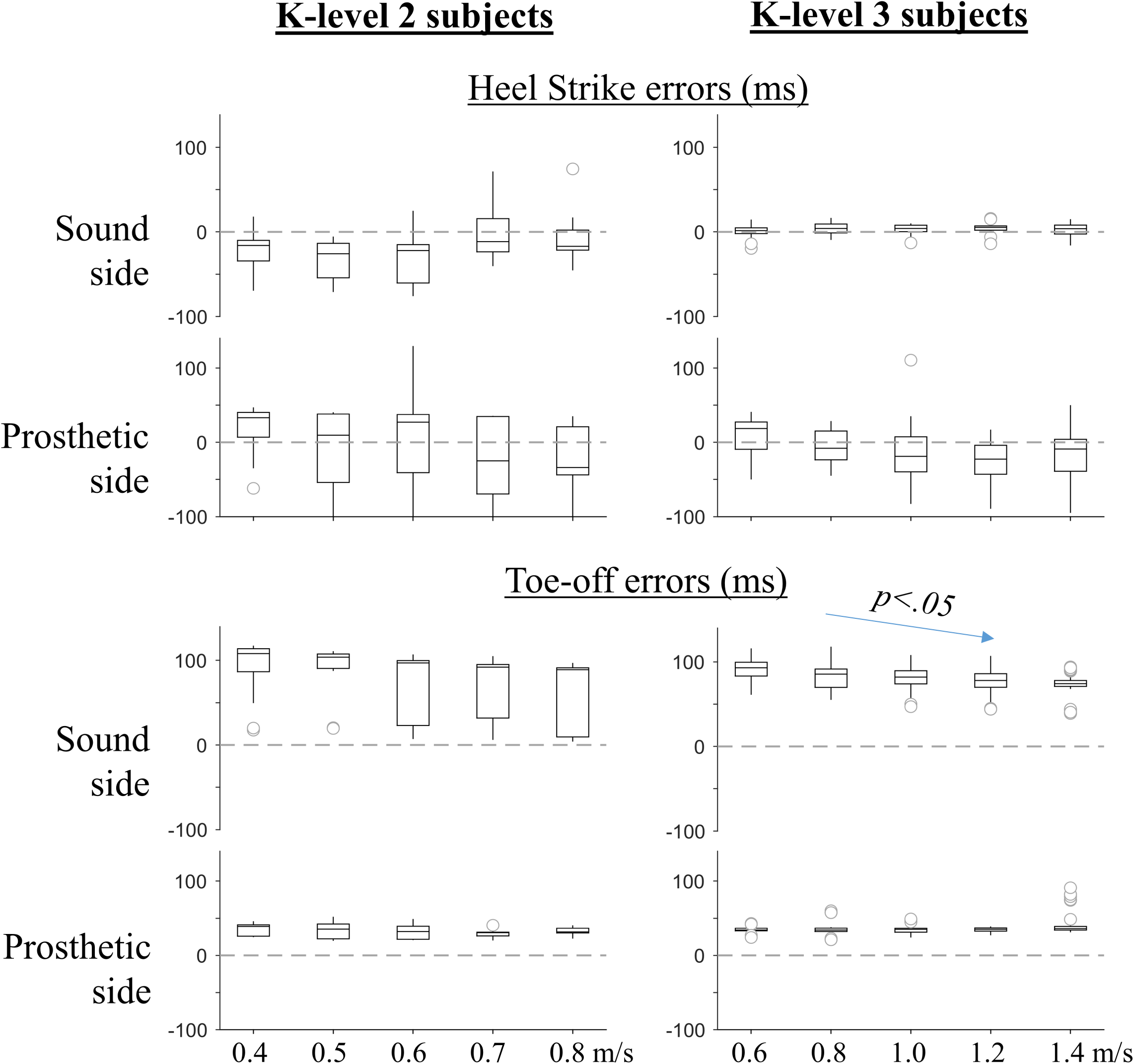
Toe-off (top panel) and Heel-strike (bottom) median error values separated by subject groups (K-level 2 on left and K-level 3 on right) and leg side. Speed is varied on x-axis for each group (0.4 to 0.8m/s for K2 subjects, 0.6 to 1.4m/s for K3 subjects)

As a whole, the K2 group results had larger dispersion compared to K3 groups indicated by larger IQR values at all speeds. Moreover, the error distribution was highly skewed for K2 subjects as indicated by asymmetrical boxplots. On the other hand, walking speed had a significant effect on the TO prediction error on the sound side only for K3 subjects (p¡.05, Friedman test). Other variables did not exhibit any significant correlation with walking speed.

## 4 Discussion and conclusion

This is one of the most comprehensive studies for any population which compares kinematics-based TO and HS prediction against the force platform data. The results provide an insight into the degree of credence of velocity-based gait event prediction. At the same time, the results can be exploited by researchers for event prediction in the absence of force platforms.

A key finding from this study is the consistent early prediction of the toe-off event for both legs. This means that the actual toe-off takes place after the 1^*st*^ negative peak or minima of the velocity signal and points towards an inherent limitation of this algorithm. In physiological terms, it indicates that the leg has already started to accelerate forward before the foot leaves the ground. Some authors have proposed the point of zero-crossing (the point where the signal crosses the from negative-to-positive) as TO event (e.g. [27]). However, observation of our velocity signals for numerous trials does not support this view. We postulate that the actual TO event occurs after the negative peak (NP) but before the zero-crossing (ZC) of the velocity signal, hence yielding a narrow NP-ZC zone. Future studies should focus on this zone for accurate prediction of TO event.

On the other hand, the HS prediction stretched both positive and negative values, indicating early and late prediction respectively. The error magnitude was however smaller indicating that the actual heel-strike occurs in a small window around the 2^*nd*^ negative peak. The result of this study can be used to define a search window around the negative peaks of the leg velocity graph as shown in Figure 5. For the HS, the window stretches about 50ms in both directions of the first negative peak. Similarly, for the TO, the actual event window stretches up to 100ms after the second negative peak. Future research should focus on these windows while improving the prediction accuracy of kinematics-based methods possibly by including information from other signals such as foot and/or leg accelerations.

**Figure 5:**
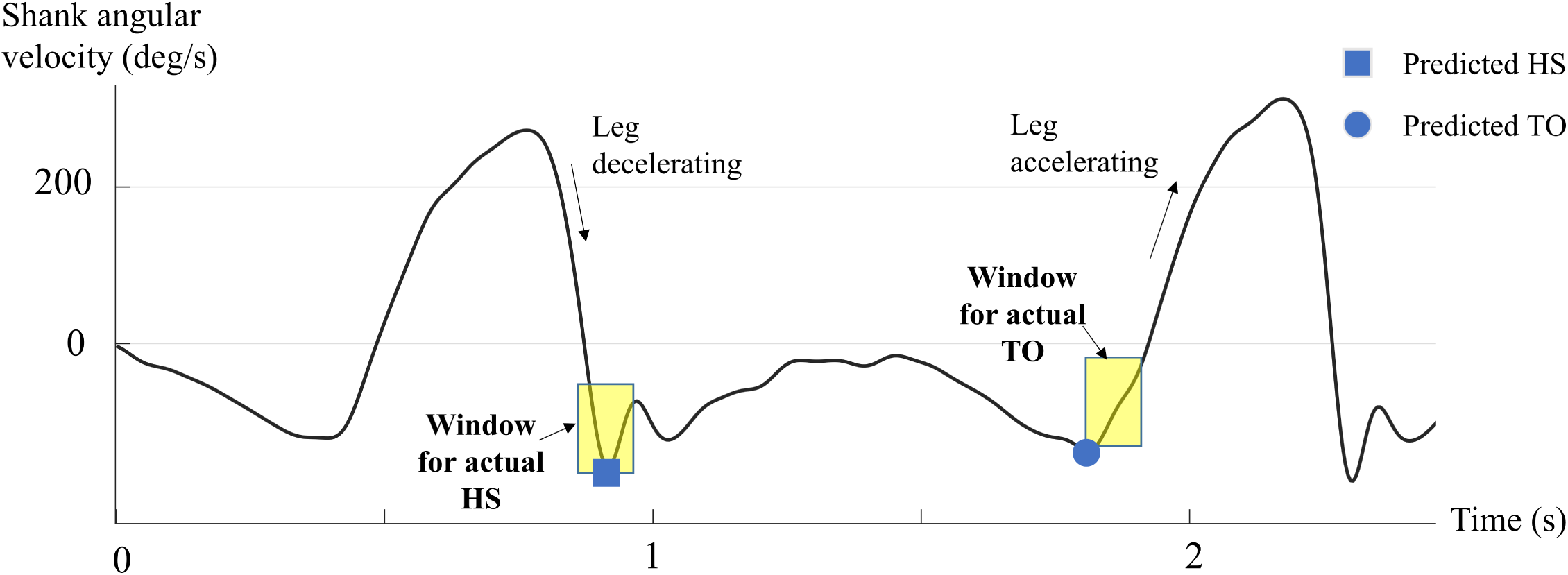
Typical shank velocity graph supplemented with the windows likely to contain the HS and TO events for a gait cycle based on the results of this study.

### Leg and group effects

Group comparisons indicated that the eTO prediction accuracy was better on the prosthetic side (Figure 3) and further correlated negatively with walking speed (Figure 4) on the sound side. On the other hand, the eHS variability was larger for K-level 2 subject group which warrants precaution when using the kinematics method for subjects classified as ‘limited community ambulators’ on the functional classification system.

### Comparison with literature studies

As mentioned in the beginning, there are barely any published studies with the amputee population which makes a direct comparison of results difficult. Nevertheless, a comparison of the results of this study with the available literature is presented in Table 2. Almost all studies have reported early TO prediction with this algorithm, albeit with smaller magnitudes than our results. Catalfamo et. al. [2] reported early TO prediction for all steps with a mean error of 50-73ms for healthy and children suffering from cerebral palsy. Trojaniello et. al. [14] reported mean absolute TO errors in the range of 16 to 22ms for elderly and gait-impaired subjects. These smaller magnitudes further reinforce our premise that the actual TO event occurs in the NP-ZC zone mentioned earlier for all populations.

**Table 2:** A comparison with the error magnitudes found in this study and the available relevant literature, ME: Mean Error, MAE: Mean Absolute Error

| Study | Subject population and task | Prediction method | HS error (ms) | TO error (ms) |
| --- | --- | --- | --- | --- |
| This study | Amputee, N=10, Level treadmill walking | Leg kinematics, Dual-minima of shank velocity | ME: -9.3, MAE: 29 | ME: 57.7, MAE: 58.6 |
| Maqbool et. al. 2015 [20] | Amputee, N=1, Ramp ascent and descent | Leg kinematics, Dual-minima | ME: -37 to 13 | ME: -17 to 122 |
| Storm et al. 2016 [3] | Healthy, N=10, Over-ground | Minima of shank velocity for HS. Acceleration-based for TO | MAE: 7 to 14 | MAE: 16 to 51 |
| Zahradka et al. 2020 [19] | Healthy and Gait-impaired, N=17, Level treadmill walking | Minima of shank velocity for TO. Zero-crossing for HS | ME: -10.45 | ME: -56.20 |
| Trojaniello et. al. 2014 [14] | Healthy and Gait-impaired, N=40, Over-ground | Minima of shank velocity for HS. Acceleration-based for TO | ME: 0 to -22, MAE 10 to 22 | ME: 0 to -16, MAE: 16 to 22 |
| Lee & Park 2011 [22] | Healthy, N=5, Overground | Leg kinematics, Dual-minima | ME: -17 to -21 | ME: 3 to 15 |
| Catalfamo et. al. 2010 [2] | Healthy and CP, N=7, Overground and ramp | Leg kinematics, Dual-minima | ME: -8 to -21, MAE: 15 to 24 | ME: 50 to 73, MAE: 50 to 73 |

Similarly, for HS prediction, the error values are smaller than for TO prediction as in this study. For instance, Zahradka et al. [19] reported a mean error of -10.45ms for a group of healthy and gait-impaired subjects which is very close to our results. Storm et. al. [3] reported absolute mean error for indoor and outdoor walking in healthy adults in the range of 11-14ms. The study by Catalfamo et. al [2] reported a mean HS error of -8ms, an absolute mean of 15ms for level-ground walking. All in all, the findings of this study match well with the literature and indicate a higher level of confidence for the HS prediction than for the TO prediction.

In conclusion, it is possible to detect heel-strike and toe-off events for the amputee population using the leg velocity, albeit less accurately for the toe-off event on the sound side and for patients with limited community ambulation ability.

## Data Availability

Available upon request

## Notes

### Competing Interest Statement

The authors have declared no competing interest.

### Funding Statement

There is no funding information to disclose

### Author Declarations

No IRB approval was required for this study.

